# Interplay of physical and cognitive performance using hierarchical continuous-time dynamic modelling and a dual-task training regime in Alzheimer’s patients

**DOI:** 10.1101/2022.12.14.22283428

**Authors:** S. Schwarck, M. C. Voelkle, A. Becke, N. Busse, W. Glanz, E. Düzel, G. Ziegler

**Affiliations:** Institute of Cognitive Neurology and Dementia research, Otto-von-Guericke-University Magdeburg, Germany; German Center of Neurodegenerative Diseases (DZNE), Magdeburg, Germany; Institute of Psychology, Humboldt University of Berlin, Berlin, Germany

**Author notes:** shared senior authorship (E. Düzel, G. Ziegler).

**Keywords:** Alzheimer’s disease, cardiovascular training, cognitive performance, intervention, continuous-time modelling, longitudinal analysis, dynamic modelling, hierarchical, Bayesian

## Abstract

**Background:** Training studies typically investigate the cumulative rather than the analytically challenging immediate effect of exercise on cognitive outcomes.

**Methods:** We investigated the dynamic interplay between single-session exercise intensity and time-locked cognition in older adults with suspected Alzheimer’s dementia (N = 17) undergoing a 24-week dual-task regime. We specified a state of the art hierarchical Bayesian continuous-time dynamic model with fully connected state variables to analyze the bidirectional effects between physical and cognitive performance over time.

**Results:** Higher physical performance was dynamically linked to improved memory recognition (-1.335, *SD* = 0.201, 95 BCI [-1.725, -0.954]). The effect was short-term, lasting up to five days (-0.368, SD = 0.05, 95 BCI [-0.479, -0.266]). Clinical scores supported the validity of the model and observed temporal dynamics.

**Conclusion:** Higher physical performance predicted improved memory recognition in a day-by-day manner, providing a proof-of-concept for the feasibility of linking exercise training and cognition in patients with Alzheimer’s dementia.

**Highlights:**

- Hierarchical Bayesian continuous-time dynamic modelling approach
- 72 repeated physical exercise (PP) and cognitive (COG) performance measurements
- PP is dynamically linked to session-to-session variability of COG
- Higher PP improved COG in subsequent sessions in subjects with Alzheimer’s dementia
- Short-term effect: lasting up to four days after training session

**Research in Context:**

1. **Systematic review:** Training-induced effects on cognitive outcomes in Alzheimer’s dementia and/or associated dynamic Bayesian modelling approaches were reviewed. Although studies showed exercise-induced cognitive improvements or maintenance, most of these studies fail to capture the dynamic nature of the change and interplay of physical (PP) and cognitive (COG) performance.
2. **Interpretation:** Using a sophisticated hierarchical Bayesian continuous-time dynamic modelling approach, a fully connected state variable model was specified. PP is dynamically linked to COG, i.e. higher PP predicted improved COG in subsequent sessions. This effect was rather short term, lasting for up to five days.
3. **Future direction:** Our results support exercise-induced effects on cognition. The cognitive system was still able to fluctuate and change favourably even in a sample with Alzheimer’s dementia. Further studies using dynamic modelling are necessary to replicate findings and examine other contributors to cognitive volatility in dementia.

## 1. Background

Alzheimer’s disease is a devastating neurodegenerative disorder encompassing neuropathological changes including accumulation of amyloid plaques, neurofibrillary tangles and neuronal as well as synaptic loss resulting in macrostructural atrophy of the brain (1). Alzheimer’s dementia is characterized by a progressive decline of functional independence in everyday life, with memory being the most affected cognitive domain (1,2). It has been shown that a higher physical fitness across the lifespan mitigates age-related cognitive decline and reduces the risk of progression to dementia (3,4). Physical fitness seems to reduce Alzheimer’s dementia -risk substantially (5) and mice- and human studies suggest a neuroprotective effect of physical activity by reducing Aβ plaques, increasing hippocampal plasticity/neurogenesis and improving memory performance (6–8). Therefore, targeted interventions focussing on physical inactivity are under investigation to delay the progression of cognitive decline (9). Several intervention-studies reported positive effects of exercise training on cognitive performance in patients with Alzheimer’s dementia or mild cognitive impairment (10). However, there is also contradictory evidence where cognition did not benefit in patients with dementia despite physical fitness improvements (11,12).

Exercise-interventions in dementia are typically envisioned as prolonged regimes of regular weekly training sessions spanning several months. During this time, the exercise intensity is gradually increased according to individual cardio-vascular and physical fitness parameters. Given the prolonged nature of exercise interventions, it is clinically relevant to understand how each training session and changing parameters of training intensity affect cognition, both during training and shortly after training. These time-locked, immediate effects of training are important for several reasons. First, they can provide mechanistic insights into the training-related day-to-day fluctuations of cognition of a patient to develop more effective approaches and guide care and clinical decision making. Second, they can provide individual guidance for adaptively choosing the optimal training intensity from a cognitive point-of-view.

To summarize, there is partial support for the hypothesis that physical fitness might positively impact cognition and counteract the cognitive decline in patients with Alzheimer’s dementia (4,5). However, the majority of studies using repeated measurements only report undirected associations rather than analysing the dynamic changes and complex interplay of physical and cognitive states directly (13). Temporal precedence is an important requirement for causality (14). Moreover, time is often treated as a discrete variable and accordingly the regression strength between time points is estimated without integrating information regarding the interval between them (14,15). Previous longitudinal studies therefore faced challenging unequal time interval lengths between and/or within participants, which might lead to biased parameter estimates and conclusions (14–16). In contrast, a hierarchical Bayesian continuous-time dynamic modelling approach overcomes problems of conventional approaches, such as above biases and lack of directionality (13,17). This approach enables the analysis of the dynamic change and mechanistic coupling between states of interest, fully accounting for unequal acquisition time intervals (14,18).

Here we study the dynamic changes of physical and cognitive performance and their interplay using an extensive longitudinal training study for patients with suspected Alzheimer’s dementia comprised of 72 sessions over 24-weeks including a physical and cognitive dual-task regime. The positive exercise-induced effect of physical fitness on cognition was shown in a previous paper using a conventional linear approach (19). In this study we focus on the dynamic interplay between physical and cognitive performance taking advantage of state of the art hierarchical Bayesian continuous-time dynamical system modelling (14,18,20). We hypothesize that changes in physical performance predict subsequent changes of cognitive performance. In addition, we hypothesize that individual dynamics relate to dementia symptom severity as well as physical health.

## 2. Methods

### 2.1 Sample and experimental design

We recruited older adults aged 60-80 years with diagnosed mild to moderate Alzheimer’s disease (F00.1; Mini Mental Status Examination (MMSE): 18-26) from the memory clinic of the German Center for Neurodegenerative Diseases (DZNE), Magdeburg (see Supplemental Material: Methods). The total sample size included N = 17 older adults (age: *M* = 73.33 ± 3.43; MMSE: *M* = 23.50 ± 3.45; female = 8) who were free of depressive symptoms (Geriatric Depression Scale: *M* = 1.88 ± 0.93), pulmonary and cardiovascular diseases. The study contained a 24-week (72 sessions, 15-min each) dual-task regime (for details see also (19)). The training support per week was provided by the relatives, a team member and a senior volunteer. Each participant received personalized training with an individualized cadence (40- 80 rotations per minute) and exercise intensity of 65-75% of the maximum heart rate (HR, ∼ 90-115 bpm) using the Karvonen method (21). Simultaneously, each training session included a cognitive task with a memorization of 30 pictures, such as animals or landscapes. The picture recognition memory performance was assessed immediately after training and included 30 subsequent test screens (original vs. lure picture). All picture sets were matched regarding difficulty level using LaMem score (22). The MMSE and 12-Item Short Form Survey (SF-12) physical health questionnaire (23) were assessed before and immediately after the regime.

### 2.2 Dynamical Bayesian modelling and statistical analysis

We included up to 72 training sessions per participant in the analysis if completed (90% in sample). The training observations were z-standardized on the group level. Physical performance (PP) was measured as the ratio of power output (unit: Watt) and HR of each training session. Higher values reflect higher PP. Cognitive performance (COG) was operationalized as reaction times corrected for the number of errors. The reaction time (RT, in seconds) within a valid range of ≤ 13 seconds was used and corrected for the proportion of error (PE) using the linear integrated speed-accuracy score (LISAS) (24):

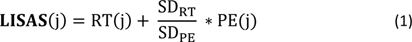

with mean reaction time and proportion of errors at the measurement occasion j, and respective standard deviations. Lower scores do reflect higher cognitive performance i.e. faster reaction time corrected for error.

All statistical analyses were conducted in R version 4.0.2 using RStudio version 1.3.1056. The modelling approach was previously established and implemented in the R package ctsem (20) relying on the Stan software (25). Results were visualized using ctsem and ggplot2 from tidyverse (26). Model fit was compared using Chi-square tests. The alpha level for additional frequentist statistical tests was defined as p < .05.

Hierarchical Bayesian continuous-time dynamic modelling was used to simultaneously analyse the temporal dynamics of PP and COG reflected in a 2-dimensional state variable **η**(t) =[PP(t), COG(t)]^T^ at time t (20). At the core is a subject-level latent dynamic model using a stochastic differential equation (or state equation):

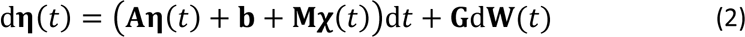

with time-varying latent process **η**(*t*) and its temporal derivative d**η**(*t*) encoding the systems current state and its change over time, respectively. The DRIFT matrix **A** contains free parameters and defines the temporal dynamics of the process with auto-effects (connectivity: main-diagonal) and cross-effects encoding interplay or interactions (coupling: off-diagonal) Eq.(2) contains the continuous time intercept (CINT) **b** and the effect **M** of time dependent predictors **χ** on **η**(*t*).

The continuous-time parameters of **A** contain changes of **η** over a small time interval in the differential equation and can be transformed better interpretable discrete-time equivalents (**A**\*) for any given time interval length (Δ*t*)

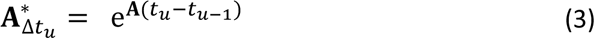

where **A**^*^_Δ*t*_*u*__ includes the associated auto and cross-regression effect for the effect of **η** at the measurement occasion *u*-1 on **η** at measurement occasion *u* (20). The used approach also includes a linear measurement model relating latent states **η**(*t*) to observables **y**(*t*)

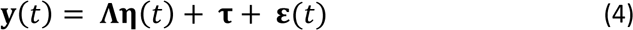

using factor loadings **Λ**, the manifest intercepts **τ** and residuals **ε** with covariance matrix **Θ** (see Supplemental Material).

The dynamic model was specified with two fully connected state variables enabling bi- directional coupling between PP and COG over 72 measurement occasions. The observable indicator Power/HR of each session were loading on PP and reaction times (RT) on COG. All other parameters (except **b** and **M** which were set to zero) of the state equation and measurement model were left free to be estimated using the data. The latent process means at t=0, **τ** (intercepts) and **A** (DRIFT-matrix) were allowed to vary free across participants, resulting in 49 parameters. Population and individual level parameters are estimated simultaneously using all data from all subjects. The hierarchical Bayesian model estimation was set to default priors and initial starting values using 4 chains and 8.000 iterations (under Stan’s optimizer for maximum a posteriori estimates).

Four models were hypothesized and further compared. First, a full 2-CR model including both auto- and cross-effect parameters in drift matrix **A** resulting in 13 free population mean parameters. Secondly, two 1-CR models with both auto-effects and the cross-effect PP on COG (PP→COG) and COG on PP (COG→PP), restricting to uni-directional interaction only. Lastly, a zero-model (0-CR) in which both cross-effects were fixed to zero (without domain interplays). The full 2-CR model was compared against both 1-CR (and 0-CR model with regard to their model fit using a Chi-Square difference test (20).

Furthermore, we ran a second-level model with the MMSE and the SF12 baseline score as time-independent covariates for validation with the temporal dynamics. A higher score in both covariates is associated with higher cognitive and physical health status respectively. All other model specifications were the same as the above full 2-CR model.

To explore potential changes of the domain interplay over the course of the training, the strength of PP on COG (PP→COG) was compared between the baseline (day 1-84) and second half (day 85-168) of the training (both free parameters to estimate). By specifying a slightly extended full 2-CR model, we included a time dependent predictor **χ** (named ‘secondhalf’), which is zero expect on day 84, when it is 1. Additional, an extra latent process (named ‘step2ndhalf’) was included, with all parameters and covariances fixed to zero, except the element on the time dependent predictor were set to 1 (step function), i.e. the extra latent process shifts on day 84 to 1 and stays there. The cross-effect in question was defined as a function of PP→COG on baseline added by PP→COG on the second half of the training multiplied by the extra latent process. A negative value reflects a stronger effect of PP→COG (i.e. when physical performance increases the reaction time decreases) in the second half of the training and vice versa.

## 3. Results

### 3.1 Demographic data

Table 1 provides the demographic data and associated neurological characteristics of the participants.

**Table 1:** Demographic data and neurological characteristics of the sample (N = 17). MMSE pre, Mini Mental Status Examination score before the start of the intervention. MMSE post, Mini Mental Status Examination score after the end of the 24-week intervention (assessment within 12 days to 8 weeks after the intervention). * N = 16 due to one missing post assessment. *M*, mean. *SD*, standard deviation.

| Demographic data |  |
| --- | --- |
| Age (years) | 67-80 ($M = 73.41$, $SD = 3.43$) |
| Sex | 7:10 (ratio female to male) |
| Neurological characteristics |  |
| ICD-10 diagnosis | Alzheimer's disease (F00.1) |
| MMSE pre | $M = 23.35$, $SD = 3.50$ |
| MMSE post* | $M = 22.75$, $SD = 4.06$ |

### 3.2 Analysis of cross-domain interplay using dynamic modelling

The range of valid measurement occasions of all 17 participants was between 59 – 70 sessions (*M* = 64.71 ± 4.26) resulting in 1100 manifest observations per latent state (or domain) with 25 (∼2.3%) missing values for physical performance. The time interval between successive measurement occasions ranged from 1 to 14 days (*M* = 2.54 ± 1.04). The longitudinal data was analysed using a 2-state dynamical model as illustrated in Figure 1 (for details see methods).

**Fig. 1.**
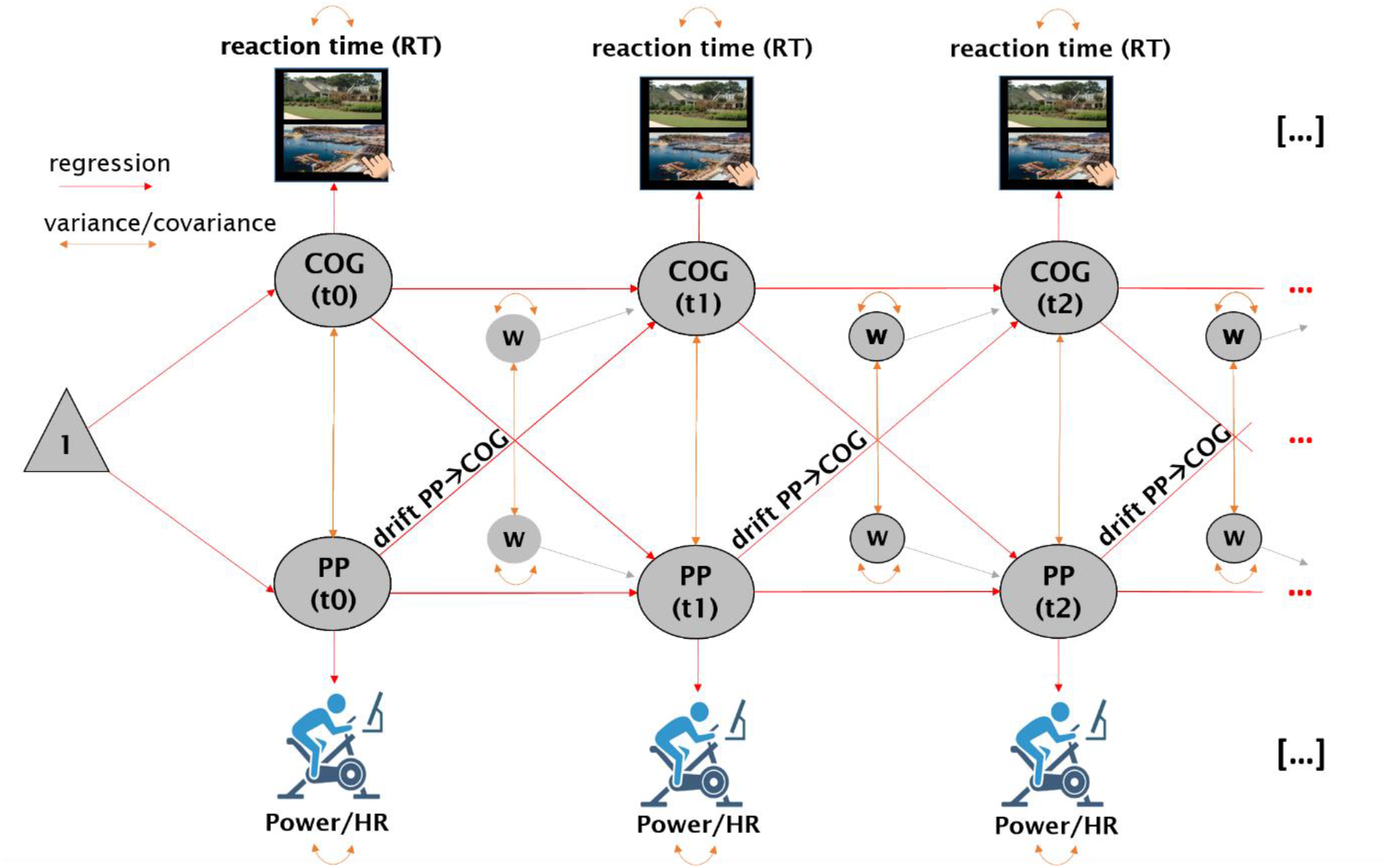
Schematic illustration of the 2-state-model with the first three timepoints (t0, t1, t2) reflecting successive training sessions. The graphical model contains the observed variables (manifest indicators) power of the bicycle ergometer and HR (as ratio: Power/HR) and reaction time corrected for the proportion of error (RT) loading on the latent variables (ellipsoids) PP (physical performance) respectively COG (cognitive performance). The main effect of interest is the cross-effect of PP on COG (further denoted as drift PP→COG). The model also contains latent error terms (**w**) and the continuous time intercept (triangle). The model shows regression paths (red lines) and variance and covariance (orange lines). Manifest intercepts are not shown.

A Chi-Square difference test for model comparison revealed a significant difference between the full 2-CR model and the zero-model (χ²(17) = 235.1, *p* < .001). In addition, a significant difference was also observed between the 2-CR model and the PP→COG 1-CR model (χ²(9) = 149.8, *p* < .001) and COG→PP 1-CR model (χ²(9) = 294.6, *p* < .001). Correspondingly, the full 2-CR model (Table 2) containing both cross-effects that enable a bi-directional interplay between COG and PP fitted the data best and is further reported (see Table S1 for estimated population parameters for the other models).

**Table 2:** Group level results showing estimated population means including Bayesian posterior intervals of the Full 2-CR (cross-effect) model. Sample size *n* = 17 with 1100 observed sessions in total; The model contains two latent variables (physical (PP) and cognitive (COG) performance) with one manifest indicator, each (Power/HR and RT corrected for PE) respectively, *n* = 13 free population mean parameters. Est. mean from mean of the chains; BCI, 95% Bayesian credible interval, LL, lower limit, UL, upper limit; Bayesian model estimation: number of chains = 4, number of iterations = 8000.

|  | *parameter* | *symbol* | *Est.* | *SD* | *LL-BCI* | *UL-BCI* |
| --- | --- | --- | --- | --- | --- | --- |
| **DRIFT** | drift<sub>PP</sub> | A | -0.851 | 0.146 | -1.146 | -0.602 |
|  | drift<sub>COG</sub> | A | -1.645 | 0.188 | -2.026 | -1.300 |
|  | drift<sub>COG→PP</sub> | A | 0.313 | 0.087 | 0.154 | 0.485 |
|  | drift<sub>PP→COG</sub> | A | -1.335 | 0.201 | -1.725 | -0.954 |
| **T0<sub>MEANS</sub>** | T0<sub>mPP</sub> | η<sub>1</sub> | -0.175 | 0.010 | -0.194 | -0.154 |
|  | T0<sub>mCOG</sub> | η<sub>2</sub> | 0.941 | 0.083 | 0.766 | 1.102 |
| **DIFFUSION** | diff<sub>PP</sub> | Q | 0.045 | 0.011 | 0.027 | 0.070 |
|  | diff<sub>COG</sub> | Q | 0.797 | 0.038 | 0.722 | 0.874 |
|  | diff<sub>PP→COG</sub> | Q | -0.687 | 0.135 | -0.884 | -0.354 |
| **MANIFEST<sub>VAR</sub>** | mVar<sub>Power/HR</sub> | Θ | 0.208 | 0.006 | 0.195 | 0.220 |
|  | mVar<sub>RT</sub> | Θ | 0.046 | 0.029 | 0.012 | 0.124 |
| **MANIFEST<sub>MEANS</sub>** | mm<sub>Power/HR</sub> | τ | -0.382 | 0.078 | -0.531 | -0.234 |
|  | mm<sub>RT</sub> | τ | 0.259 | 0.079 | 0.103 | 0.412 |

Changes in PP predict later changes in COG in the opposite direction as found by a more substantial negative cross-effect drift_PP◊COG_ (-1.335, *SD* = 0.201, 95 BCI [-1.725, -0.954]). As such, when physical performance levels are above baseline (suggesting higher PP) COG levels are likely to go downwards (faster reaction times). The cross-effect PP→COG also varies between participants which suggests that some subjects benefit more from the exercise training resulting in cognitive improvements than others (between-person variability in drift_PP◊COG_: 2.44, *SD* = 0.29, 95% BCI[1.86, 3.02]). The high between-person differences in the cross-effect PP→COG comes along with its higher population mean (T0_MEANS_, Table 2), i.e. this effect seems more relevant in the sample but its importance also differs more between participants. In contrast, COG do partially predict later changes of PP in the same direction as indicated by a small and positive cross-effect (drift_COG◊PP_). When subject’s reaction time increases (i.e. reduced COG) the PP levels did also slightly increase over following time. Furthermore, the temporal changes of PP last longer than the temporal dynamic of COG (higher auto-effect of PP (drift_PP_)).

Previous states of PP do impact COG negatively (-0.368, *SD* = 0.05, 95 BCI [-0.479, -0.266]) as shown via discrete-time parameters (one day time interval). The expected effect PP→COG peaks around one and lasts for up to around four days, after which the random-state fluctuations dominate (Figure 2). The cross-lagged effect COG→PP was observed to be close to zero (0.086, *SD* = 0.024, 95 BCI [0.041, 0.136]) and accordingly there is practically no substantial effect in this direction. Furthermore, the small autoregressive (self-connection) effect of COG (0.145, *SD* = 0.039, 95 BCI [0.072, 0.226] suggested low stability of the construct over time. PP on one day had a small effect on PP on another day (0.363, *SD* = 0.05, 95 BCI [0.264, 0.472].

**Fig 2.**
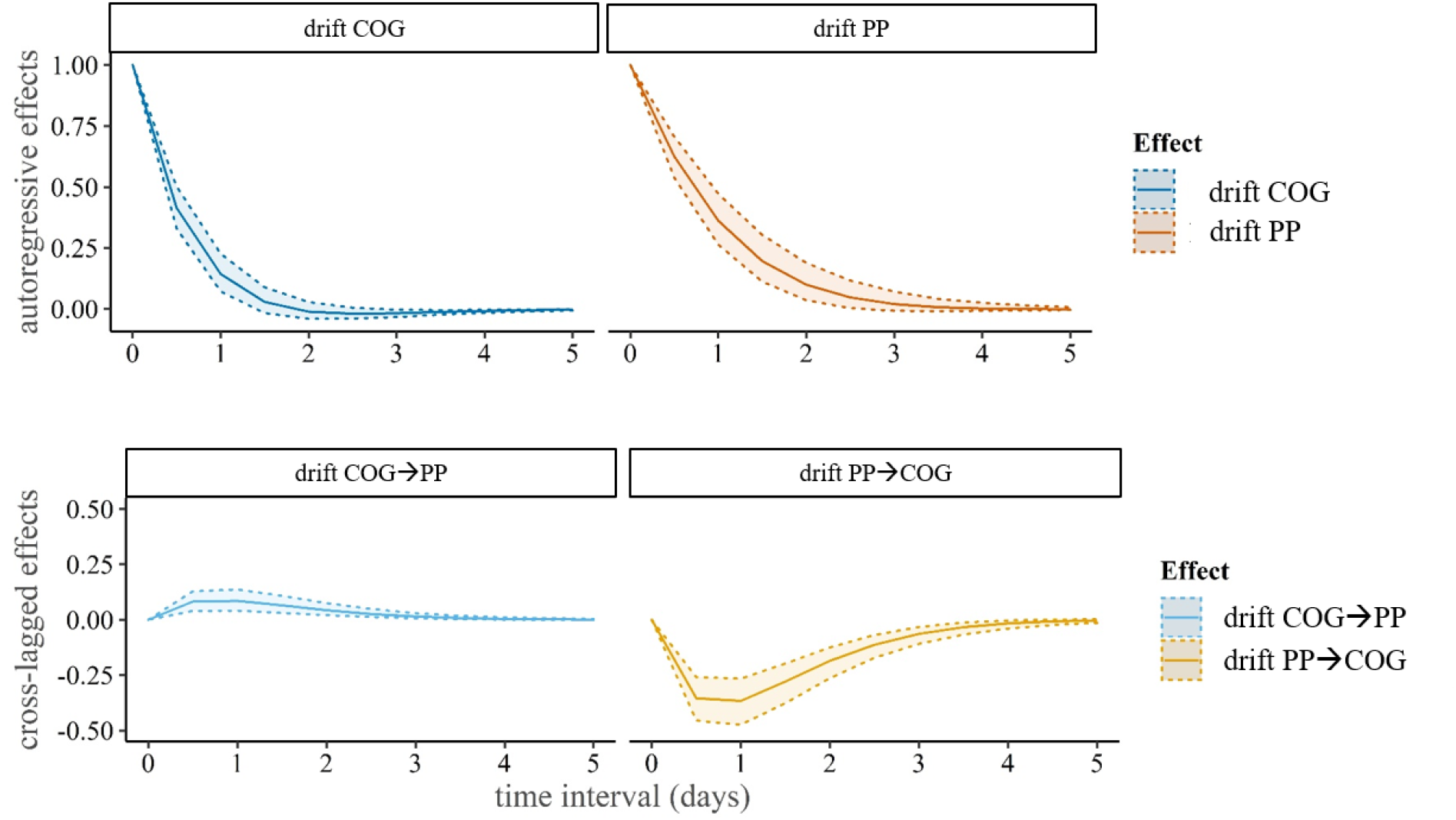
Auto and cross regression over time. Temporal autoregressive effects (upper panel) and cross-lagged effects (lower panel) over time (x-axis, time interval in days), median and 95% quantiles for a change of 1 at time zero. The expected autoregressive effect (or self-connection) of PP (drift PP) and COG (drift COG) peak around approximately one day and decrease with increasing time interval length. This suggests that the more time passes the less predictive is the performance for consecutive performance levels. The expected cross-lagged effect (or interplay) of PP on COG (PP→COG) peaks around one day and seem to improve predictions of COG for up to around four days. This can be understood as rather short-term benefits from physical training on cognitive performance. The cross-lagged effect COG on PP (COG→PP) is very close to zero, suggesting that changes in cognitive performance do not improve predictions of physical performance.

The temporal dynamics of PP showed more inter-individual differences compared to COG. Some subjects showed relatively persistent PP levels over the entire training time, while the PP fluctuated more in other participants (between-person variability in drift_PP_: 9.89, *SD* = 0.52, 95% BCI[8.89, 10.90]). The measurement error (MANIFEST_VAR_) of the manifest indicator Power/HR (0.208) was found to be higher compared to RT (0.046), i.e. measurement limitations and short-term situational influences (e.g. subjective stress or sunny days) are more present in the PP indicator (Table 2)., COG showed higher session-to-session fluctuations within-person (0.188, *SD* = 0.014, 95 BCI [0.163, 0.217]) compared to PP (0.004, SD = 0.002, 95 BCI [0.001, 0.012]). The model prediction of COG and PP over training time are illustrated in Figure 3 (see also Supplemental Material Fig. S1 for five randomly selected participants).

**Fig. 3.**
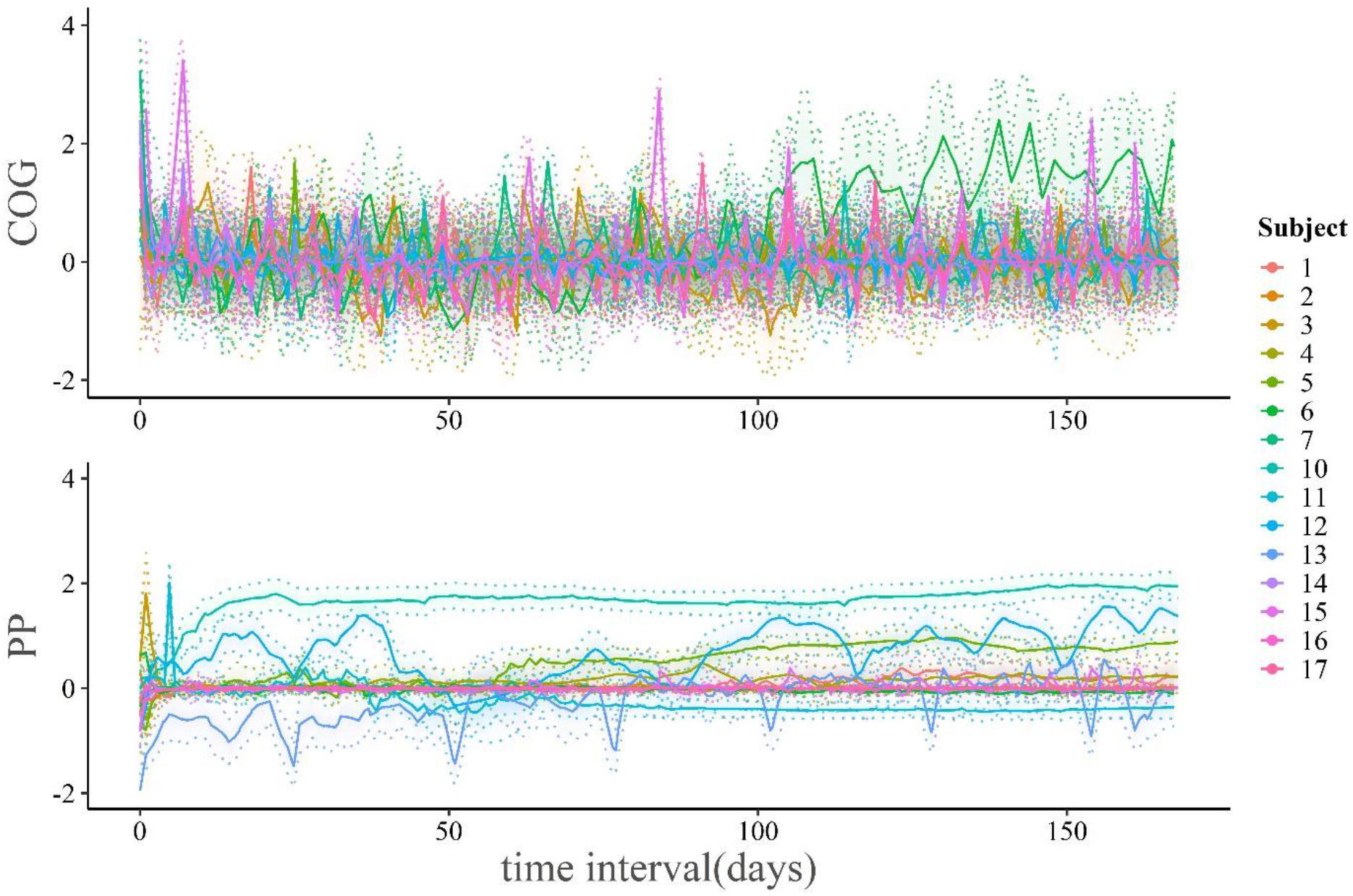
Individual estimates. Individual level analyses for all participants of the sample (*n* = 17) over time interval in days (x-axis). The solid lines presents the model prediction of the smoothed estimates of participant’s individual latent states COG (upper panel) and PP (lower panel) within a 95% BCI. Each coloured solid line presents the individual model prediction for one subject each. The temporal dynamics of PP show more individual differences compared to COG.

### 3.3 Drift coefficients as a function of clinical baseline scores

Participants with higher MMSE baseline scores show lower persistence in the cross-effect COG→PP (drift_COG◊PP_; -0.215, *SD* = 0.022, 95% BCI [-0.259, -0.172], Fig 4A). Higher MMSE baseline scores were associated with lower persistence in COG (drift_COG_; -0.538, *SD* = 0.174, 95% BCI [-0.887, -0.209]). The effect of MMSE on the auto-effect PP and the cross-effect PP→COG is close to zero (-0.02, *SD* = 0.003, 95% BCI [-0.03, -0.01]. Participants with a lower health score show a stronger cross-effect of COG on PP (drift_COG◊PP_; -0.231, *SD* = 0.035, 95% BCI [-0.303, -0.165], Fig 4B). Likewise, higher physical health scores seem to be associated with lower persistence in their physical performance (drift_PP_; -0.036, *SD* = 0.008, 95% BCI [- 0.052, -0.021]). The effect on the other auto-effect and cross-effect are close to zero.

**Fig. 4.**
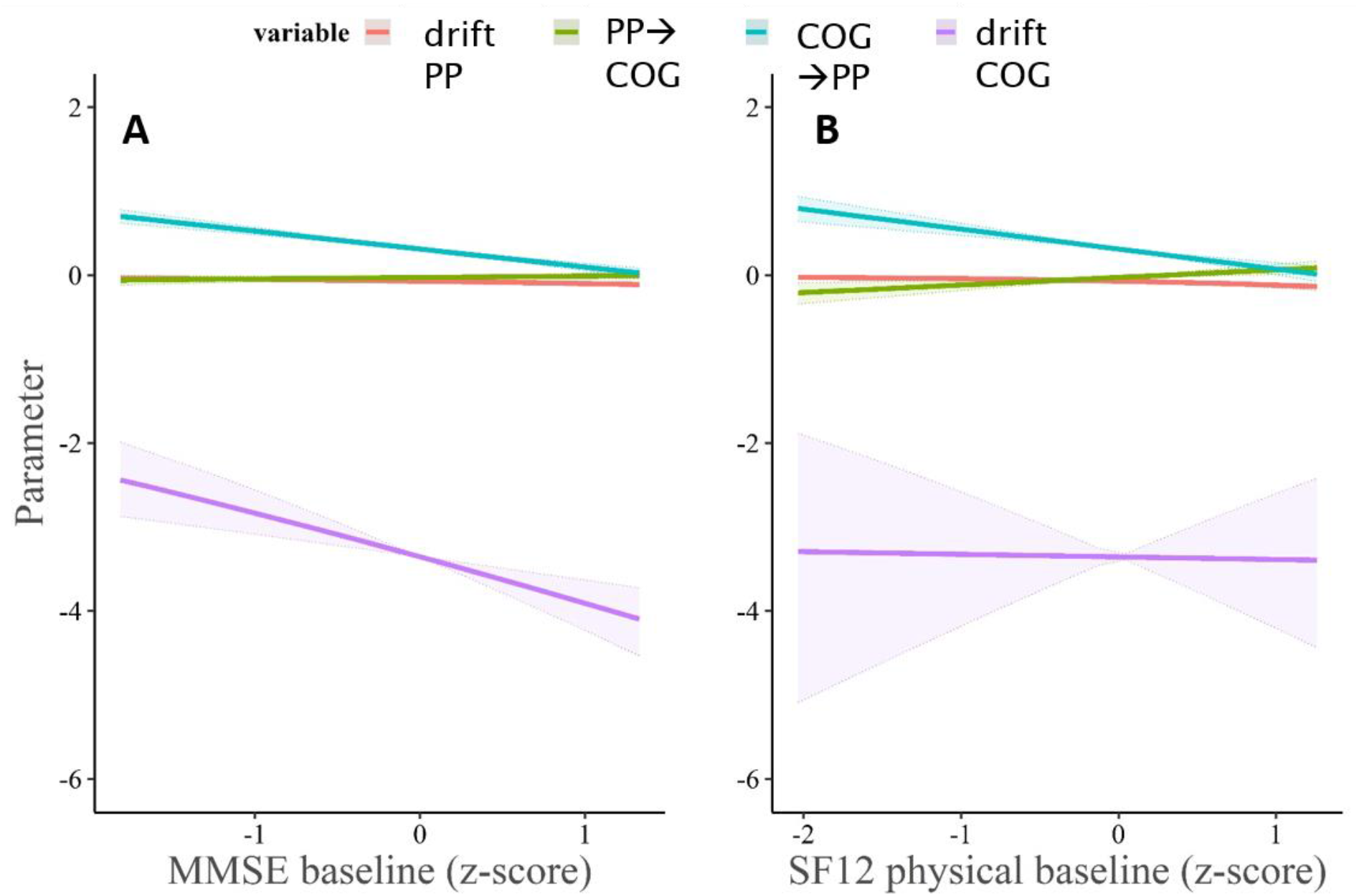
Estimated effect of subject-level covariate predictors on dynamical parameters. We show (A) MMSE and (B) SF12 physical health baseline score effects on drift parameters (auto-effects and cross-effects) within a 95% BCI. drift_PP_, auto-effect PP (red solid line); drift_PP◊COG,_ cross-effect PP on COG (green solid line); drift_COG◊PP,_ cross-effect COG on PP (blue solid line); drift_COG,_ auto-effect COG (purple solid line).

### 3.4 Changes of dynamics over the course of training

An extended model with the drift coefficient PP→COG as a function of time was specified. The model estimated if the strength of the cross-effect PP→COG changed between the baseline (day 1-84) and the second half (day 85-168) of training. Results suggested that the cross-effect PP→COG becomes positive in the second half of the training (2.08), i.e. the strength and associated effect of physical performance on cognitive performance was found to be reduced in the second half of training. This change was estimated as a function of PP→COG on baseline (-0.11, *SD* = 0.63, 95% BCI [-1.34, 1.14]) added PP→COG on the second half of the training (-1.50, *SD* = 0.08, 95% BCI [-1.66, -1.34]) multiplied by the extra latent step process (-1.45, SD = 0.38, 95% BCI [-2.21, -0.72]).

## 4. Discussion

Recent longitudinal studies reported promising but also conflicting effects of exercise training on cognitive performance in persons with dementia (10). These studies often fail to capture the dynamic nature of the coupling between fitness and cognition (14). This study assessed the dynamic session-to-session changes and interplay of physical and cognitive performance by taking advantage of a Bayesian hierarchical continuous-time dynamic modelling approach (20). A fully connected model was specified to analyse the connectivity (auto/self-effect) and coupling (cross-domain/interplay) of PP and COG over 72 measurement occasions.

Under the assumption of our model, physical performance is dynamically linked to cognitive performance. Thus, increased PP was associated with improved COG (faster reaction time), which is in line with previous training studies (4,5). However, this effect shows between-person variability, suggesting that some participants benefited more from the training than others. While we cannot rule out the possibility of non-responders due to a missing control group (27), the observed cross-effect supports the notion that exercise training has a positive effect on cognition in dementia.

In addition, an important question is how long this beneficial exercise-induced effect on cognition lasts (e.g. how many days). Our results suggest rather a short-term effect: A positive change in PP can improve the prediction of increased COG for up to four days with the strongest influence after one day, after which unpredictable random fluctuations dominate. However, unexplained or not modelled causes could act on this effect and might appear even without training. Nevertheless, the coupling-effect appears at least under the condition of training, consistent with recent studies (4,5).

Furthermore, we observed random session-to-session cognitive fluctuations within participants over time, while the measurement error was close to zero. In patients with Alzheimer’s dementia, intra-individual cognitive fluctuations are generally higher compared to healthy controls (28,29) and may be linked to pathology (30). Considering this fluctuations as measurement error only would oversimplify the true cognitive state (31).

The observed short-term effects in patients with dementia suggests that exercise seem to mobilize capacities that would otherwise remain unused. However, the exercise-induced causal underlying mechanism(s) leading to cognitive changes are still discussed (32,33). There is some evidence for neurotrophin-mediated neurogenesis, such as BDNF (brain-derived neurotrophic factor) (34,35). Inhibited BDNF in the hippocampus by blocking tyrosine receptor kinase B also inhibited the action of BDNF and its associated beneficial effects on cognitive function (36). The short-term nature of the effect that we report here suggests that rather than the chronic and slow plasticity related to neurogenesis, more immediate mechanisms may play a role. These include changes in the bodily milieu, improved perfusion and improved clearance, just to name a few (35,37,38).

We further support some validity for our observed temporal dynamics by assessing the drift coefficients as a function of clinical baseline scores. Higher physical health (SF-12) and lower cognitive impairment (MMSE) were associated with lower persistence in PP and COG, respectively.

PP showed a general increasing trend and COG a decreasing trend (faster reaction times) over training time. This is in line with our previous study using a linear-mixed modelling approach (19). Beyond, we observed changes in the coupling-effect of PP on COG over training time by specifying an additional model. The coupling-effect was stronger in the first half of training (day 1- 84), which is line with training studies (39,40). However, in the second half of training (day 85 – 168) the strength of this coupling-effect is reduced, i.e. the exercise-induced increase of PP positively affecting COG appeared to weaken over time. One explanation might be that the changes may follow a non-linear time course, e.g. increase early or close to the end of the whole training regime. As such, further studies using time-varying analysis of those domains may clarify the temporal patterns of the coupling-effect. However, patients with Alzheimer’s dementia show general reduced motivational capacities (41). Animal studies observed positive effects of environmental enrichment (combined physical, cognitive and social stimulation) on brain health, such as increased neurotrophic factor levels, neurogenesis, and improved cognitive performance (42). Although our training was designed as a cognitive motivation, i.e. the participants had to physically exercise for new pictures to stimulate the novelty-exploration, the lower variability in the task itself may have led to reduced motivation and/or increased distraction (e.g., by social or environmental factors) from the middle of the training onwards.

Finally, we like to mention several limitations of the current study. Although the model is mechanistic (or causal) it contains many assumptions that might be wrong and/or we may have not included all observable factors mediating the observed effects. Our approach assumed stationary dynamics over the course of the training, i.e. we cannot rule out non-stationary dynamics during the training time. The sample was small (N=17) and potentially biased regarding age and severity of Alzheimer’s dementia and did not include a control group. Since there is an absence of a physically inactive control group, future studies might look if cognitive volatility (i.e. also short reaching performance highs) is present without training and whether the coupling of physical on cognitive performance still exist without. With regard to the present literature showing positive due to regular exercise (3,4), the latter one might not be the case. Future longitudinal randomized controlled studies using a more comprehensive neuropsychological assessment to examine transfer effects are necessary to verify our results. Additionally, studies may examine the question how cognitive fluctuations can be used since they are random.

The present extensive 24-week longitudinal training study examined the temporal connectivity und coupling of physical and cognitive performance in a sample with Alzheimer’s dementia using a hierarchical Bayesian continuous-time dynamic modelling approach. Physical performance was dynamically linked to cognitive performance, i.e. higher PP improved time- locked COG in subsequent sessions. The beneficial effect was rather short-term, lasting up for four days after the training. Our observed dynamics were validated by clinical scores, i.e. higher MMSE baseline scores were associated with lower persistence in the temporal dynamics of COG. To summarize, our results provided a proof-of-concept regarding the feasibility of time-resolved linkage of exercise training and cognition even in a sample with suspected Alzheimer’s dementia.

## List of abbreviations

BDNF: brain-derived neurotrophic factor
COG: cognitive performance
CR: cross-effect
HR: heart rate
HRV: heart rate variability
LISAS: linear speed-accuracy score
MCI: mild cognitive impairment
MMSE: Mini Mental Status Examination
PE: proportion of error
PP: physical performance
RT: reaction time
SF-12: 12-Item Short Form Survey
SF-36: 36-Item Short Form Survey

## Declarations

### Ethics approval and consent to participate

The study was approved by the ethics committee of the Otto-von-Guericke University, Magdeburg, Germany (approval number: 68/17). All participants and their relatives as representatives signed a written informed consent form for participation.

### Availability of data and materials

The datasets used and/or analysed during the current study are available from the corresponding author on reasonable request.

### Competing interests

The authors declare that they have no competing interests.

### Funding

The project (ID: ZS/2016/05/78611) belongs to the Research Association Autonomy in old Age (AiA) funded by the European Union (ERDF-European Regional Development Fund) and the State of Saxony-Anhalt, Germany.

### Author’s contributions

GZ and SS contributed to the conceptualization and the data curation. SS, GZ, and MV performed the formal analysis. ED contributed to the funding acquisition, resources, and the supervision. The investigation was conducted by NB, WG, AB, and SS. ED, MV, GZ, AB, and SS contributed to the methodology and AB and NB to the project administration. The software and validation was conducted by AB. SS contributed to the visualization and the writing of the original draft preparation. SS, GZ, MV, WG, AB, NB, and ED contributed to the writing of the review and editing. All authors contributed to the article and read and approved the submitted version.

## Data Availability

All data produced in the present study are available upon reasonable request to the authors

## Acknowledgements

Not applicable

## Notes

### Competing Interest Statement

The authors have declared no competing interest.

### Clinical Trial

DRKS registration number: DRKS00019105

### Author Declarations

The ethics committee of the Otto-von-Guericke University, Magdeburg, Germany gave ethical approval for this work (approval number: 68/17).

### Summary of Updates

Manuscript was shortened; Part of Methods are now included in in the Supplemental Material

