## Supplementary material for "Interplay of physical and cognitive performance using hierarchical continuous-time dynamic modelling and a dual-task training regime in Alzheimer’s patients": DADM_TS1_ctsem_final_Supplements_20230807.pdf

### Supplemental Material

#### Methods

##### Sample and experimental design

The recruitment for the study took place in the period from September 2017 to September 2018 from the memory clinic of the German Center for Neurodegenerative Diseases (DZNE), Magdeburg. During enrolment 125 patients were assessed for eligibility. 92 patients were excluded because they did not meet the inclusion criteria (n = 10), declined to participate (n = 65) and due to other reasons (n = 17) such as too small apartments for an ergometer. In addition, 14 participants did not receive the allocated intervention due to deterioration of symptoms. One participant dropped out during the intervention due to physical health difficulties.

All participants and their relatives as representatives signed a written informed consent form for participation. The study was approved by the ethics committee of the Otto-von-Guericke University, Magdeburg, Germany (approval number: 68/17). The study was registered as a clinical trial after the enrolment of participants started since this study was planned as a feasibility study (DRKS registration number: DRKS00019105). The authors confirm that all ongoing and related trials for this intervention are registered. The participants were not compensated monetarily for the costs of participation.

The dual task regime used in this study is a simultaneous physical and cognitive training, which is specifically developed for older adults with Alzheimer's dementia. It was attempted that each participant exercised at the same time to better control circadian rhythm. The dual task regime took place in the own households of the participants.

Each training session began with the individual start-resistance and proceeded by successively increasing it each 60s until the individual target heart rate (HR) was reached. Moreover, the overall training regime included an increase of the resistance from the individual assessed start resistance by approximately 5% every four weeks. The subjective perceived exertion was assessed after each session using a 6-20 Borg Scale. These individual adaptations were carried out if the average target heart rate and the ratings of perceived exertion after each training session were in the determined range. The HR was continuously measured every second using a Garmin chest belt. The pictures were presented one after the other for 20 seconds each on an integrated tablet (Samsung Galaxy Tab A 2016 10.1 inch). During each 10-second inter-stimulus interval the screen background turned white. The same set of pictures was presented during each of the three training sessions within one week while the applied set was varied over weeks to minimize learning effects. The different sets of pictures were matched regarding the average difficulty memorability level determined by the open source LaMem score (Khosla et al., 2015).

Finally, a picture recognition test was carried out to measure visual short-term memory performance. The test was designed without a time limit for the response. The pictures on each test screen were arranged on top of each other, with the arrangement of the original and lure picture randomised. The lure picture was new but matched the memorability score of the original picture. The picture recognition memory test was designed as a forced choice task. The participants had to decide one by one which of both test pictures was the original. As with the pictures visible during the dual task, each training week contained the same lure pictures, while the 24 picture sets per week differed from each other. The lure picture sets were also matched for memorability using the LaMem score (Khosla et al., 2015).

#### Dynamical Bayesian modelling and statistical analysis

Subject-level latent dynamic model

$$\mathrm{d}\boldsymbol{\eta}(t) = \left(\mathbf{A}\boldsymbol{\eta}(t) + \mathbf{b} + \mathbf{M}\boldsymbol{\chi}(t)\right)\mathrm{d}t + \mathbf{G}\mathrm{d}\mathbf{W}(t) \tag{2}$$

Furthermore, $\mathbf{W}(t)$ denotes the so-called Wiener process, a random walk in continuous time with covariance matrix $\mathbf{Q} = \mathbf{G}\mathbf{G}^{\mathrm{T}}$. $\mathbf{Q}$ is also referred as the diffusion matrix. Further technical details and explanations can be found in (58).

Thirdly, the framework allows for some (or potentially all) parameters to differ across individuals. Here the parameters for each subject are drawn from a population distribution with unknown mean and variance and priors. Additionally, effects of time independent predictors (covariate effects) can be included (see Driver and Voelkle (2018) for further details).

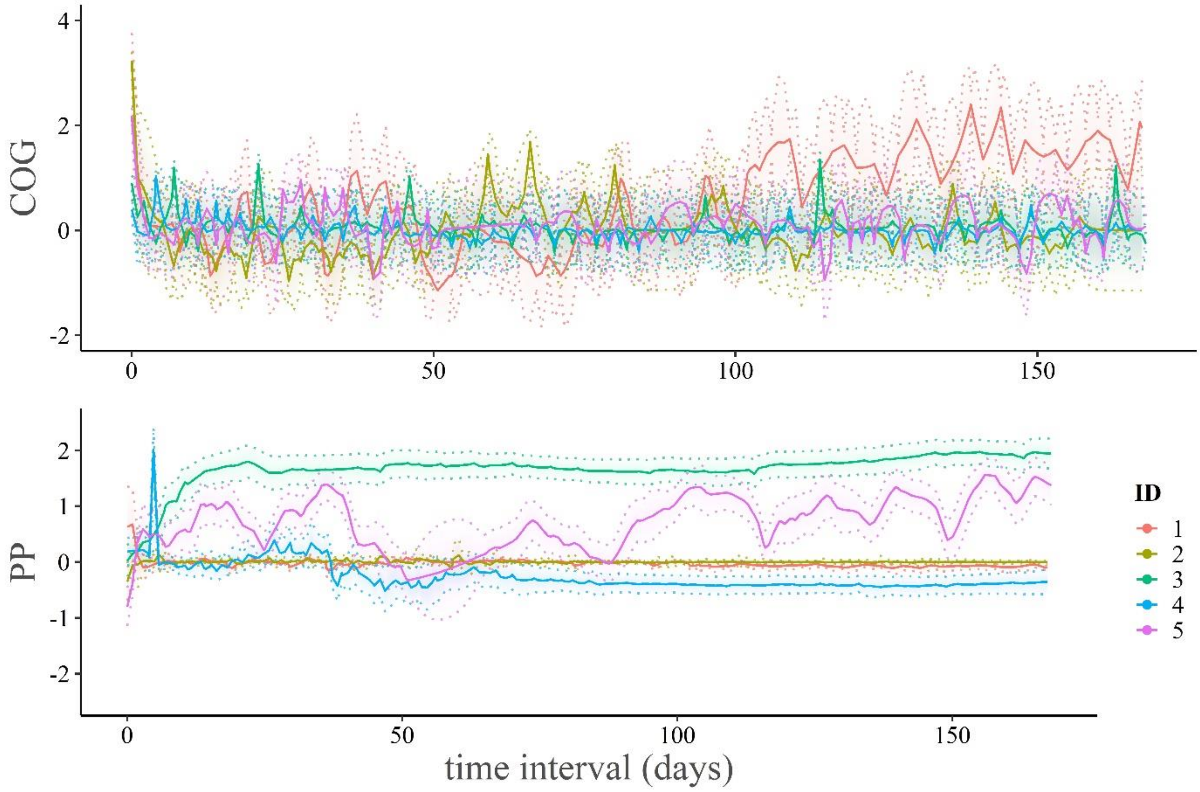


Fig. S1. Individual level analyses for five randomly selected participants of the sample over time interval in days (x-axis). The solid lines presents the model prediction of the smoothed estimates of participant's individual latent states COG (upper panel) and PP (lower panel) within a 95% BCI. Each coloured solid line presents the individual model prediction for one subject (ID) each. The temporal dynamics of PP show more individual differences compared to COG.

Alternative model examine dynamic change and interplay of physical (PP) and cognitive (COG) performance

| | | | *model with cross-effect COG→PP only* | | | | *model with cross-effect PP→COG only* | | | | *model with no cross-effects* | | | |
|---|---|---|---|---|---|---|---|---|---|---|---|---|---|---|
| | parameter | symbol | **Est.** | **SD** | **LL-BCI** | **UL-BCI** | **Est.** | **SD** | **LL-BCI** | **UL-BCI** | **Est.** | **SD** | **LL-BCI** | **UL-BCI** |
| DRIFT | drift$_{PP}$ | A | -0.00 | 0.01 | -0.01 | -0.00 | -0.50 | 0.17 | -0.90 | -0.24 | -4.42 | 0.75 | -5.92 | -2.96 |
| | drift$_{COG}$ | A | -5.89 | 0.84 | -7.53 | -4.31 | -3.43 | 1.81 | 7.52 | -0.73 | -1.74 | 0.36 | -2.49 | -1.09 |
| | drift$_{COG \rightarrow PP}$ | A | 0.48 | 0.09 | 0.30 | 0.66 | | | | | | | | |
| | drift$_{PP \rightarrow COG}$ | A | | | | | -0.52 | 0.29 | -1.09 | 0.04 | | | | |
| T0$_{MEANS}$ | T0m$_{PP}$ | $\eta_1$ | -0.15 | 0.11 | -0.36 | 0.06 | -0.31 | 0.02 | -0.36 | -0.25 | -0.18 | 0.05 | -0.28 | -0.08 |
| | T0m$_{COG}$ | $\eta_1$ | 0.30 | 0.05 | 0.18 | 0.41 | 0.95 | 0.63 | -0.18 | 2.15 | 1.11 | 0.30 | 0.47 | 1.67 |
| DIFFUSION | diff$_{PP}$ | Q | 0.03 | 0.01 | 0.02 | 0.05 | 0.33 | 0.02 | 0.30 | 0.38 | 0.17 | 0.02 | 0.13 | 0.21 |
| | diff$_{PP \rightarrow COG}$ | Q | -0.69 | 0.12 | -0.86 | -0.41 | 0.03 | 0.05 | -0.73 | 1.34 | -0.08 | 0.06 | -0.16 | -0.01 |
| | diff$_{COG}$ | Q | 1.07 | 0.15 | 0.80 | 1.39 | 1.00 | 0.16 | 0.73 | 1.34 | 0.82 | 0.06 | 0.70 | 0.96 |
| MANIFEST VAR | mvar$_{Power/HR}$ | $\Theta$ | 0.20 | 0.01 | 0.18 | 0.21 | 0.03 | 0.02 | 0.01 | 0.11 | 0.24 | 0.01 | 0.22 | 0.25 |
| | mvar$_{RT}$ | $\Theta$ | 0.24 | 0.03 | 0.18 | 0.32 | 0.10 | 0.07 | 0.02 | 0.30 | 0.07 | 0.04 | 0.02 | 0.18 |
| MANIFEST MEANS | mm$_{Power/HR}$ | $\tau$ | -0.38 | 0.14 | -0.65 | -0.11 | -0.22 | 0.16 | -0.55 | 0.09 | -0.29 | 0.17 | -0.62 | 0.06 |
| | mm$_{RT}$ | $\tau$ | 0.82 | 0.22 | 0.38 | 1.24 | 0.32 | 0.37 | -0.34 | 1.05 | 0.03 | 0.09 | -0.14 | 0.03 |

Table S1: Group level results showing estimated population means including Bayesian posterior intervals of the alternative models that were compared with the full 2-CR (cross-effect) main model. Sample size *n* = 17. All alternative models contain two latent variables (physical (PP) and cognitive (COG) performance) with one manifest indicator, each (Power/HR and RT corrected for PE). The model with cross-effect COG→PP only contains both auto-effects and the cross-effect cognitive on physical performance only, *n* = X free population mean parameters. The model with cross-effect PP→COG contains both auto-effects and the cross-effect physical on cognitive performance only, *n* = X free population mean parameters. The model with no cross-effect contains both auto-effects only, *n* = X free population mean parameters. Est. mean from mean of the chains; BCI, 95% Bayesian credible interval, LL, lower limit, UL, upper limit; Bayesian model estimation: number of chains = 4, number of iterations = 8000.
